# Translation, adaptation, and measurement properties of the Muscle-Strengthening Exercise Questionnaire among university students in Indonesia

**DOI:** 10.1101/2024.05.01.24306693

**Authors:** Reza Setyono Ashari, Rahmaningsih Mara Sabirin, Stevani Tia Bella Merlinda, Dilia Ananda Pratiwi, Melati Nilna Tsania, Rakhmat Ari Wibowo

## Abstract

**Background:** Despite the abundant evidence showing the benefits of muscle-strengthening exercise (MSE), no epidemiological tool is available for assessing MSE among Indonesian university students. This study aimed to adapt the Muscle-Strengthening Exercise Questionnaire (MSEQ) into the Indonesian context and test the validity and reliability of the MSEQ Indonesian version.

**Methods:** Translation and cultural adaptation, content validity studies, concurrent validity studies, and reliability studies were conducted with a total of 121 respondents. The concurrent validity study compared the results of measuring MSE frequency, intensity, duration, and volume with those of the 7-day diary and relative handgrip strength. For the reliability study, the respondents were asked to fill the MSEQ Indonesian version twice with a time interval of 7 days.

**Results:** Moderate-to-strong correlations were observed between the results for the weekly frequency, duration, intensity, and volume of MSE and those of the 7-day diary and hand grip strength. Test–retest reliabilities were good to excellent for machine weight, holistic, and overall MSE but poor for bodyweight MSE. In assessing the target muscle group, the MSEQ Indonesian version showed good test–retest reliability for machine-weight MSE but poor-to-very good test–retest reliability for bodyweight, free weight, and holistic MSE.

**Conclusions:** Our study showed acceptable validity and reliability of the MSEQ Indonesian version to be used for assessing MSE among university students in Indonesia. Further studies are warranted to ensure the generalizability of our findings among the adult population.

## Introduction

Recent guidelines strongly recommend adults to engage in muscle-strengthening physical activities (MSPAs) at least twice a week in addition to moderate–vigorous aerobic physical activity (MVPA) to acquire additional health benefits^[1]^. Muscle-strengthening exercise (MSE) is a form of MSPAs that uses weight training equipment, elastic bands, free weights, or body weight and is performed during leisure time^[2]^. Compared with the MSPAs that are accrued during occupational or domestic activities and may increase the risks of musculoskeletal problems, MSE is backed up by consistent evidence showing its health benefits, including lowering the risk of mortality^[3]^.

No surveillance system comprehensively covering the important dimensions of MSPAs is available in many countries, including Indonesia^[4],[5]^. The absolute burden associated with the lack of physical activity is very high in middle-income countries, including Indonesia, because of their large population size^[6]^. Despite the potential benefit of physical activity in reducing noncommunicable diseases^[7]^, the number of people undertaking sufficient physical activity in Indonesia has declined in the recent 5-year period (2013 to 2018)^[8]^. Furthermore, the university student population, a key population for Indonesia’s demographic dividend, performs less physical activity than the general adult population. Given the health, cognitive, and academic outcome benefits of supplementing MSPAs to MVPA^[9]^, the establishment of MSPA surveillance systems among university students in Indonesia should be advocated.

The Muscle-Strengthening Exercise Questionnaire (MSEQ) is a valid and reliable instrument designed to assess MSE in an online, self-report format developed for the adult population^[10]^. The items assessed include the following dimensions: 1) frequency, 2) intensity, 3) duration, 4) type of exercise, and 5) targeted muscle groups of MSE^[11]^. However, the MSEQ is still unavailable in Bahasa Indonesia and has not been considered in the Indonesian context. Therefore, our study aimed to adapt and translate the MSEQ to Bahasa Indonesia. A pilot test was conducted to examine its validity and reliability among university students.

## Methods

This study was part of a larger study on the implementation and evaluation of a physical activity program in a university setting. We conducted this study in accordance with the Declaration of Helsinki after obtaining an ethical approval from the Medical and Health Research Ethics Committee Faculty of Medicine, Public Health, and Nursing Universitas Gadjah Mada (Ref No: KE/FK/0661/EC). Written informed consent from participants was obtained and documented before data collection.

The MSEQ was translated to Bahasa Indonesia and culturally adapted to the Indonesian context by employing Beaton’s and Sousa’s guidelines and a previous questionnaire translation study (Figure 1)^[11],[12],[13]^. The initial Indonesian version of the questionnaire was revised according to the results of content validation and then pilot-tested to examine the concurrent validity and reliability of the final version among university students and to report dropout rates and other problems that arose. Owing to the limited evidence on the gold standard for assessing the frequency, intensity, type, duration, and target muscle group of MSE in free-living contexts, concurrent validity was assessed rather than criterion validity to determine the extent of its agreement with the other noncriterion measures of MSE^[14],[15]^.

**Figure 1.**
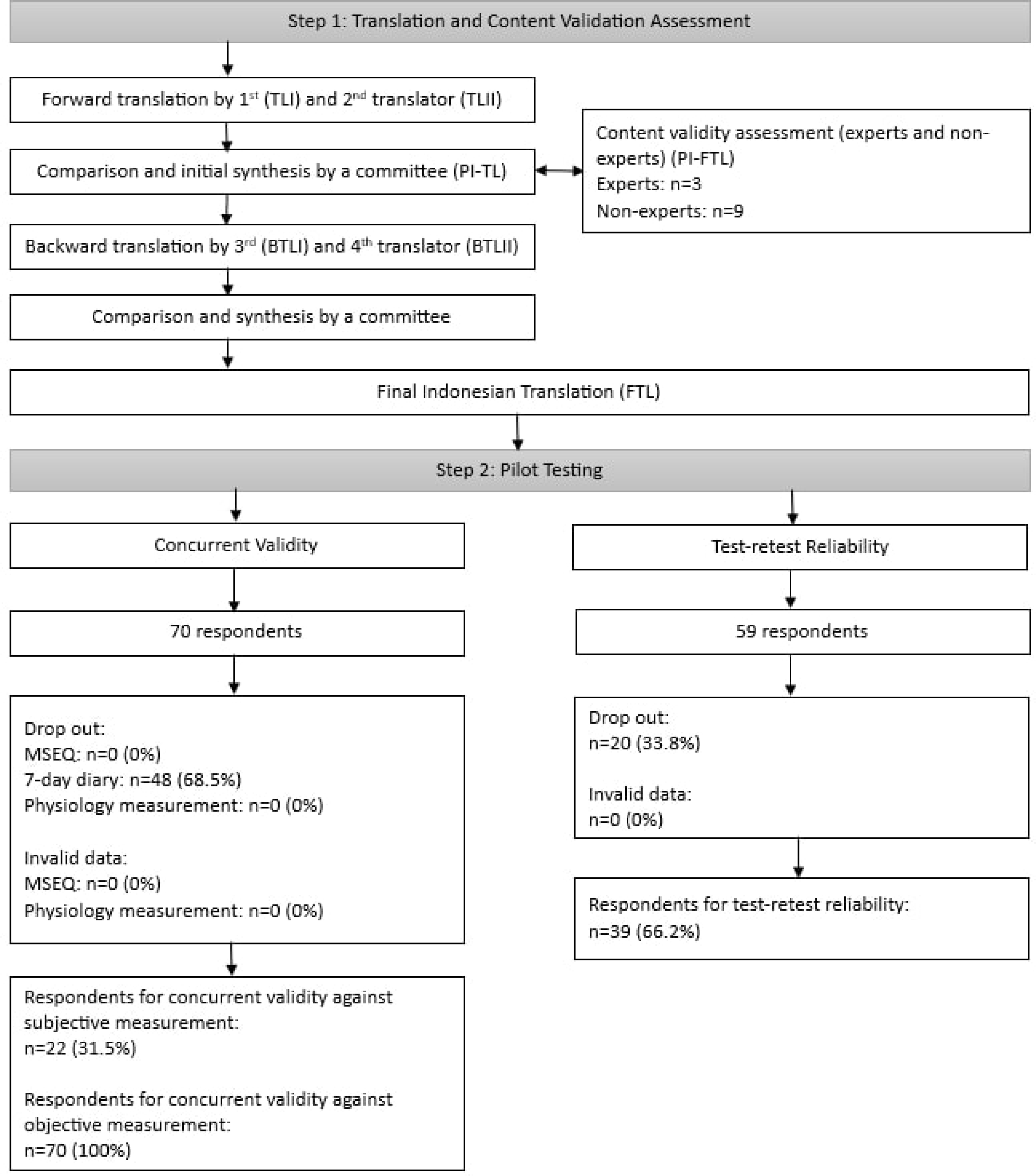

### Translation

Forward translations were conducted by two independent translators to translate the English/original version to Bahasa Indonesia, producing translation I (TL I) and translation II (TL II). The 1st translator is an Indonesian who has excellent English proficiency and is naïve in the physical activity field, and the 2nd translator is an Indonesian who has excellent English proficiency and a master’s degree in the physical activity field. A committee consisting of four researchers covering three academic fields (physical activity for health, medicine, and physiotherapy) compared TL I and TL II with the English/original version in the aspects of completeness, appropriateness, and comprehension and synthesized a preliminary initial translated version (PI-TL). The committee then assessed the content validity of PI-TL by interviewing experts and discussing with nonexpert informants. On the basis of the results, the committee discussed and revised PI-TL into a prefinal version of the translated version (P-FTL). Backward translations were independently conducted by the 3rd and 4th translators, producing back translation version I (BTL I) and back translation version II (BTL II), respectively. The 3rd translator is a UK English native who is naïve to the physical activity field and has lived in Indonesia for more than 20 years, and the 4th translator is an Indonesian who has lived in the UK for more than 10 years. Finally, a committee compared BTL 1 and BTL II to the original version, synthesized a final translated version (FTL), and created guidelines to score and analyze the MSEQ Indonesian version (Supplementary material 1). The FTL was then tested for concurrent validity and reliability.

### Validity study

#### Content validity

Content validity assessments using a mixed method approach were conducted during the translation and cross-cultural adaptation to assess the completeness, appropriateness, and comprehension of PI-TL. Purposive sampling was conducted from 4^th^ July 2022 to recruit experts (a public health researcher, an exercise science researcher, and a practitioner) and nonexperts representing university students. Guided by interview and discussion guides (Supplementary material 2), interviews with experts and FGDs with nine university students were conducted. All interviews and group discussions were audio recorded and then transcribed verbatim. Two researchers analyzed the verbatim transcripts of the interviews and discussions using the framework method^[16]^. In addition, a survey using the COnsensus-based Standards for the selection of health status Measurement Instruments (COSMIN) questionnaire for assessing content validity^[17]^ (Supplementary material 3) was administered to the experts and nonexpert informants before the interview or group discussion to examine the content validity of PI-TL. The total score was 50 points, representing 25 points of completeness, 5 points of relevance, and 20 points of comprehension. The expert and nonexpert informants were then asked to assess the completeness, appropriateness, and comprehension of P-FTL using a similar questionnaire for content validity assessment.

#### Concurrent validity

Concurrent validity was assessed by comparing the final version of the Indonesian MSEQ to a 7-day diary as a subjective criterion measure and handgrip strength (HGS) as an objective criterion measure. The 7-day diary was chosen because of its short recall period, resulting in high accuracy in capturing behaviors^[18]^. Meanwhile, HGS was chosen because it represents a physiological measure of the MSE’s habit effect^[19]^. For the pilot study, 66 respondents were set as the minimum sample population to adequately assess and identify problems to anticipate 10.6% inattentive responses^[20],[21]^. Students were conveniently recruited from a public university in Java Island, Indonesia from 22^nd^ August to 4^th^ September 2022. HGS was assessed on their dominant hand in the morning following the Southampton protocol using a hand dynamometer (Camry EH101, Zhongshan Camry Electronic, China)^[22]^. The age- and sex-specific z-score of BMI-adjusted absolute maximal HGS was used to consider the age-, sex-, and BMI-dependent HGS normative data^[23],[24],[25],[26]^. On the same day before the HGS assessment, the respondents were asked to fill out the self-administered MSEQ Indonesian online form, followed by the 7-day diary for 7 consecutive days. The researchers provided a daily reminder for the respondents. The respondents who failed to fill the diary in 24 hours were categorized as dropouts.

### Reliability study

A minimum sample of 73 respondents was set to adequately examine the test–retest reliability of the MSEQ Indonesian version by anticipating 20% dropout and 10.6% inattentive response^[20]^. A convenient sampling was conducted in one public and one private universities on Java Island and a public university on Sumatra Island through social media from 24^th^ October to 6^th^ November 2022. The respondents were asked to fill out the self-administered MSEQ Indonesian version online form twice at a 7-day interval.

### Statistical analysis

Data were analyzed using the Statistical Package for the Social Sciences V.22 (SPSS, IBM). Correlation between MSEQ and 7-day diary was examined using Spearman rank correlation and categorized as strong (at least 0.8), moderately strong (0.6–0.8), fair (0.3–0.5), or poor (<0.3)^[27]^. Bland–Altman plots were also created to report the mean of the difference and precision for the agreement between the questionnaire and the diary^[28]^. The mean of the difference below half of twice/week of MSPAs was considered acceptable as suggested by the guidelines^[1]^. Partial correlation tests controlling for BMI were conducted between the HGS z-score and the weekly frequency, weekly duration, average weekly intensity, and total weekly volume of MSE to examine the concurrent validity of the MSEQ Indonesian version to the physiological outcomes of MSE. Calculations were conducted for the intraclass coefficient correlation (ICC) of the weekly frequency, duration, volume, and average intensity of MSE and the Cohen’s kappa and agreement of the target muscle group captured using the MSEQ Indonesian version. ICC values less than 0.5 were categorized as poor, 0.5 to 0.75 as moderate, 0.75 to 0.9 as good, and greater than 0.90 as excellent reliability^[29]^. Cohen’s kappa less than 0.2 was categorized as poor, 0.21 to 0.4 as fair, 0.41 to 0.6 as moderate, 0.61 to 0.8 as good, and 0.81 to 1 as very good^[30]^.

## Results

Our study flow and characteristics of respondents are summarized in Figure 1 and Table 1.

**Table 1.** Respondents’ Characteristics.

|  |  | **Content**<br>**Validity** | **Concurrent Validity** |  | **Reliability**<br>**(n=39)** |
| --- | --- | --- | --- | --- | --- |
|  |  | **FGD sources**<br>**(n=9)** | **Subjective**<br>**(n=22)** | **Objective**<br>**(n=70)** |  |
| Age (mean±SD)<br>in years |  |  |  |  |  |
| **Sex** | Male (n[%]) | 2 (22.2%) | 2 (9.1%) | 9 (12.8%) | 13 (26.7%) |
|  | Female (n[%]) | 7 (77.8%) | 20 (90.9%) | 61 (87.2%) | 88 (73.3%) |
| **Level of study** | Undergraduate<br>(n[%]) | 5 (55.6%) | 21 (95.4%) | 69 (98.5%) | (71.1%) |
|  | Postgraduate (n[%]) | 4 (44.4%) | 1 (4.6%) | 1 (1.5%) | 13 (28.9%) |
| **Field of study** | Social sciences<br>(n[%]) | 3 (33.3%) | - | 6 (8.6%) | 23 (51.1%) |
|  | Health sciences<br>(n[%]) | 5 (55.6%) | 22 (100%) | 61 (87.1%) | 13 (28.9%) |
|  | Natural sciences<br>(n[%]) | 1 (11.1%) | - | 3 (4.3%) | 9 (20%) |
| **Type of**<br>**residence** | Private rent (n[%]) | 6 (66.7%) | 14 (63.7%) | 40 (57.1%) | 31 (68.9%) |
|  | University dormitory<br>(n[%]) | 1 (11.1%) | 1 (4.6%) | 7 (10%) | 2 (4.4%) |
|  | With parents (n[%]) | 2 (22.2%) | 7 (31.7%) | 23 (32.9%) | 12 (26.7%) |

### Content validity

We made several adjustments to the questionnaire, including adding and adjusting illustrations; customizing the options; adding day options to frequency questions; explaining intensity, exercise type, and duration; adding pages; adjusting layout; and emphasizing some explanations by applying bold marks (Supplementary material 4). Before the adjustments, the questionnaire got a score of 40.89 points, consisting of 20.11 points for relevance, 4.11 points for comprehensiveness, and 16.67 points for comprehension. Points for relevance (20.56 points), comprehensiveness (4.22 points), and comprehensibility (17.56 points) increased after the adjustment, resulting in a total of 42.34 points.

### Subjective concurrent validity

Data from 22 out of the 71 recruited respondents were analyzed to examine the concurrent validity of the MSEQ Indonesian version against the 7-day diary. However, 49 respondents dropped out from the 7-day diary recording, leaving only 22 respondents for analysis. Most of the respondents in the final analysis were female (90.9%), undergraduate students (95.4%), and lived in a private-rented house (63.7%) (Table 1).

All the MSE dimensions had a strong correlation (r range 0.73 to 0.82) (Table 2). Bland–Altman plots showed that the Indonesian MSEQ can be considered acceptable to measure all the MSE dimensions, with a slight overestimation of duration and intensity and underestimation of volume at +0.09 minutes/week, +0.15 MET, and −56.95 MET minutes/week, respectively (Figure 2).

**Figure 2.**
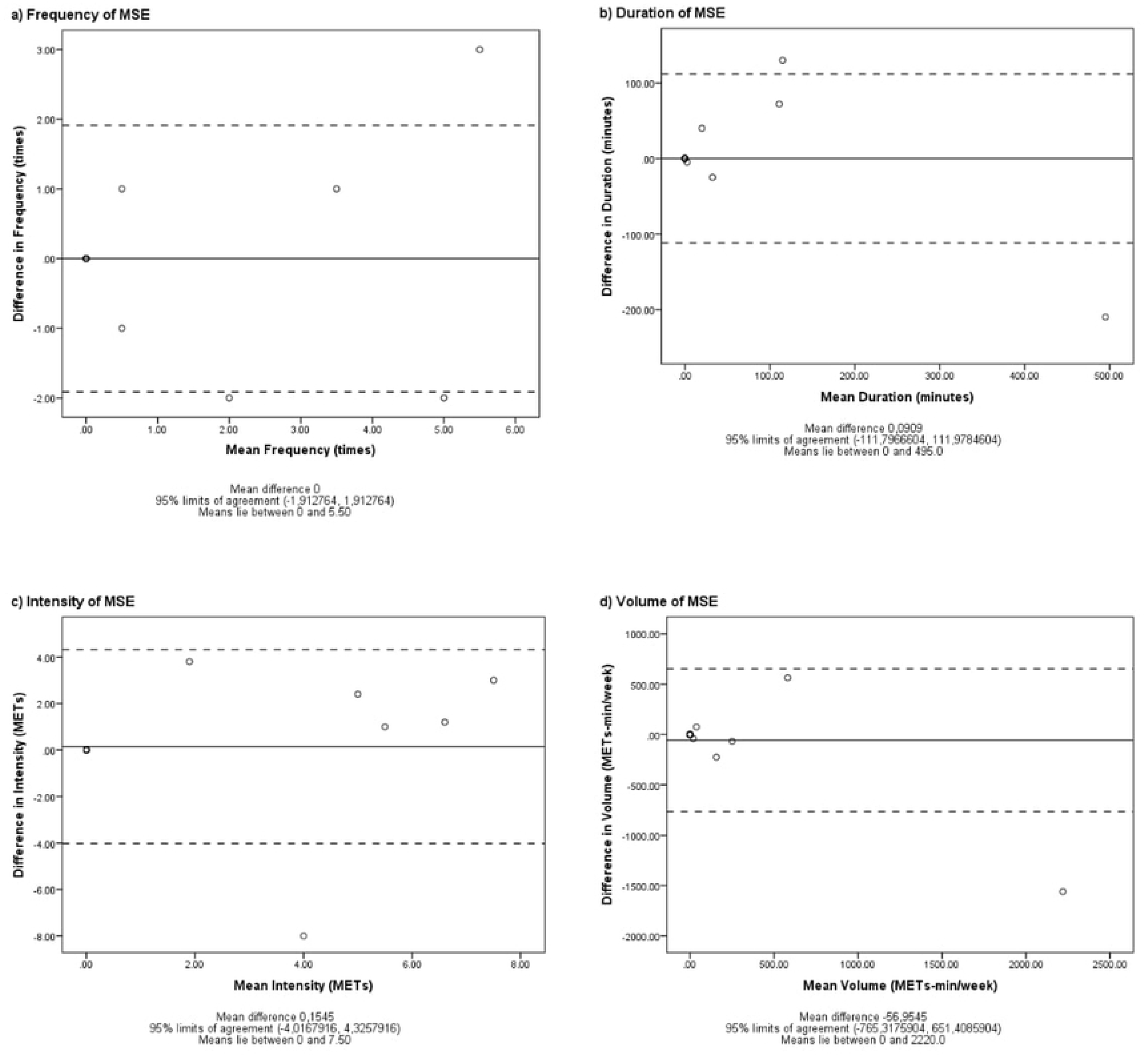

**Table 2.** Spearman Test Relative 7-Day Diary and HGS to MSE frequency, duration, average intensity, and volume of MSEQ.

| Measurement | Dimension of MSE | Spearman rho | P value |
| --- | --- | --- | --- |
| 7-Day Diary | Frequency | 0.82 (95% CI 0.60-0.92) | 0.000* |
|  | Duration | 0.81 (95% CI 0.58-0.92) | 0.000* |
|  | Average intensity | 0.73 (95% CI 0.45-0.88) | 0.000* |
|  | Volume | 0.81 (95% CI 0.59-0.92) | 0.000* |
| Hand-grip strength | Frequency | 0.35 (95% CI 0.25-0.77) | 0.003* |
|  | Duration | 0.30 (95% CI 0.12-0.54) | 0.011* |
|  | Average Intensity | 0.32 (95% CI 0.07-0.50) | 0.006* |
|  | Volume | 0.35 (95% CI 0.10-0.52) | 0.003* |

\*Statistically significant at 0.05 level of significance.
MSE, Muscle-Strengthening Exercise; MSEQ, Muscle-Strengthening Exercise Questionnaire

### Objective concurrent validity study

Data from 70 out of the 71 respondents were examined to examine the concurrent validities of MSE reported through the MSEQ Indonesian version against the relative maximal HGS as physiological outcomes (Figure 1). One respondent dropped out. Most of the respondents in the final analysis were female (87.2%), undergraduate students (98.5%), and lived in a private-rented house (57.1%) (Table 1). A moderate correlation (r range from 0.30 to 0.35) was observed between the relative maximum HGS and the frequency, duration, average intensity, and volume of MSE in the MSEQ (Table 2).

### Reliability study

The MSEQ Indonesian version showed moderate-to-excellent test–retest reliability (ICC range from 0.54 to 0.99) in assessing the frequency, duration, intensity, and volume of overall, machine weight, free weight, and holistic MSE. Its reliability was poor in examining body weight MSE (ICC range from 0.23 to 0.48) for all dimensions (Table 3) and poor-to-very good (Cohen’s kappa range from −0.13 to 0.79) in assessing the target muscle groups of body weight, free weight, and holistic MSE (Table 4). Meanwhile, it had very good reliability in assessing the target muscle groups of machine-weight MSE (Cohen’s kappa range from 0.84 to 1) (Table 4).

**Table 3.**
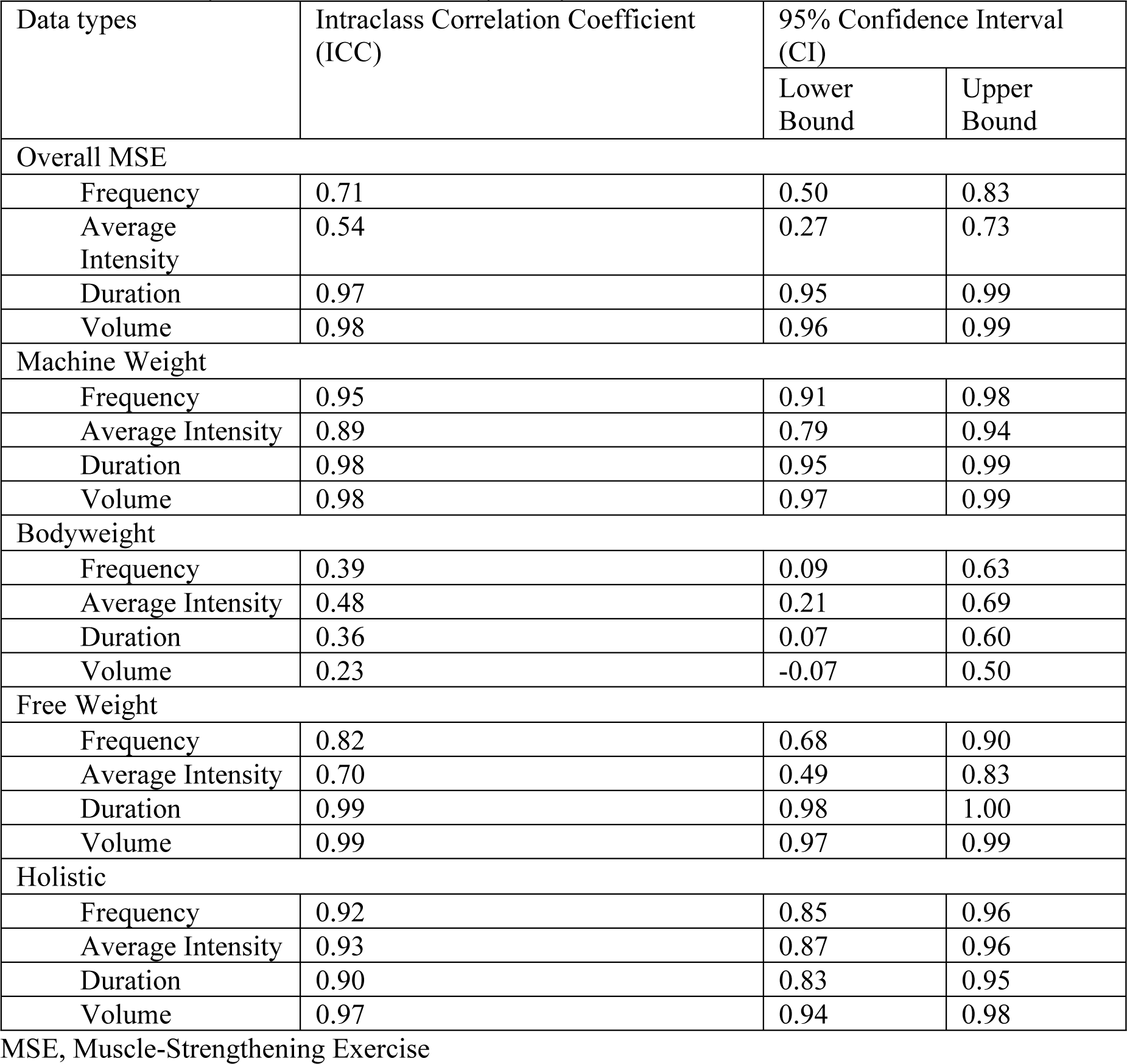
ICC Analysis Results of Reliability Study.

**Table 4.** Cohen’s Kappa Analysis Result of Reliability Study.

| Domains | Cohen's Kappa Coefficient<br>(SE of Kappa) | 95% Confidence<br>Interval (CI) |  | Percentage<br>Agreement (%) |
| --- | --- | --- | --- | --- |
|  |  | Lower<br>Limit | Upper<br>Limit |  |
| Machine Weight |  |  |  |  |
| Legs | 1 (0) | 1 | 1 | 100 |
| Hips | 1 (0) | 1 | 1 | 100 |
| Back | 1 (0) | 1 | 1 | 100 |
| Abdomen | 1 (0) | 1 | 1 | 100 |
| Chest | 0.84 (0.15) | 0.54 | 0.14 | 97.4 |
| Shoulders | 0.84 (0.15) | 0.54 | 0.14 | 97.4 |
| Arms | 0.84 (0.15) | 0.54 | 0.14 | 97.4 |
| Bodyweight |  |  |  |  |
| Legs | 0.66 (0.13) | 0.41 | 0.90 | 84.6 |
| Hips | 0.36 (0.19) | -0.01 | 0.73 | 82.05 |
| Back | -0.13 (0.06) | -0.24 | -0.01 | 71.8 |
| Abdomen | 0.47 (0.15) | 0.18 | 0.76 | 76.9 |
| Chest | 0.36 (0.19) | -0.03 | -0.74 | 82.05 |
| Shoulders | -0.11 (0.05) | -0.21 | -0.02 | 76.9 |
| Arms | 0.27 (0.16) | -0.04 | 0.58 | 79.5 |
| Free Weight |  |  |  |  |
| Legs | 0.79 (0.21) | 0.38 | 1.19 | 97.4 |
| Hips | 0.47 (0.32) | -0.15 | 1.09 | 94.9 |
| Back | 0.66 (0.32) | 0.03 | 1.28 | 97.4 |
| Abdomen | -0.04 (0.03) | -0.08 | 0.01 | 92.3 |
| Chest | 0.47 (0.32) | -0.15 | 1.09 | 94.9 |
| Shoulders | -0.07 (0.03) | -0.13 | 0.00 | 87.2 |
| Arms | 0.64 (0.24) | 0.18 | 1.10 | 94.9 |
| Holistic |  |  |  |  |
| Legs | - | - | - | 97.4 |
| Hips | 0.64 (0.24) | 0.18 | 1.10 | 94.9 |
| Back | -0.03 (0.02) | -0.06 | 0.01 | 94.9 |
| Abdomen | 0.64 (0.23) | 0.19 | 1.09 | 94.9 |
| Chest | 0.66 (0.32) | 0.03 | 1.28 | 97.4 |
| Shoulders | - | - | - | 97.4 |
| Arms | 0.48 (0.31) | -0.12 | 1.08 | 94.9 |

SE, Standard Error

## Discussion

Our study reported the translation and adaptation of the MSEQ into its Indonesian version. We have made several modifications based on themes developed by end-users’ and experts’ opinions. Owing to the translation and cultural adaptation, an improvement was observed from 40.89 to 42.34 of the 50 score of the COSMIN quantitative questionnaire for assessing content validity^[17],[31]^. In this pilot study, the MSEQ Indonesian version showed strong correlation and good precision with the 7-day diary but a weak correlation with HGS. It also had acceptable test–retest reliability in assessing the frequency, intensity, duration, and volume of MSE. However, it only showed acceptable reliability in assessing the targeted muscle groups of machine-weight MSE.

Compared with the original version, the MSEQ Indonesian version had higher test–retest reliability (ICC range from 0.51 to 0.96 and from 0.54 to 0.97, respectively) and concurrent validity (ρ range from 0.30 to 0.77 and from 0.73 to 0.82, respectively) in assessing the frequency, duration, and intensity of overall MSE^[10]^. This finding indicated that the addition of definition and explanation questions enhanced the comprehension of the MSEQ Indonesian version and consequently its reliability and validity^[32]^. Although they failed to improve the validity of GPAQ in assessing the intensity of aerobic activity^[33]^, the show cards illustrating MSE examples might have helped the subjects identify the type of MSE they engaged in because MSE might be harder to understand than aerobic activities^[34]^. The provided list of days in the option to recall the frequency of MSE functioned as memory cues that help the subjects recall their behavior; this phenomenon can also explain the improvement in the validity and reliability of the MSEQ Indonesian version^[35]^. Meanwhile, the addition of several components helped the subjects identify and recall their MSE behavior but also resulted in a high burden for them, leading to loss of interest and an increase in nonresponse items^[36],[37],[38],[39],[40]^. Our study found a 1.4% dropout rate for a single administration of MSEQ and a 33.8% dropout rate for two consecutive administrations of MSEQ at a 7-day interval. Future studies using the MSEQ Indonesian version are recommended to use reminders and prenotification to maintain the completion rate^[38],[39]^.

The MSEQ Indonesian version’s test–retest reliability score had an almost similar pattern to the original version in assessing target muscle groups. It also had an almost perfect agreement in assessing target muscle groups during MSE using weight machines (Cohen’s kappa range from 0.84 to 1). Several experts argued that MSE using weight machines is easier and does not require complex knowledge compared with the other MSE domains^[41]^. Therefore, users may find it easier to recall their MSE using weight machines compared with the other MSE domains.

In addition to the MSEQ, several questionnaires have recently been developed to assess MSE, including the 2015 Behavioral Risk Factor Surveillance System (BRFSS) for assessing the weekly frequency of MSE^[42]^, the Cancer Prevention Study-3 (CPS-3) for assessing the weekly duration of MSE^[43]^, and the Muscle-Strengthening Activity Scale (MSAS) for assessing the frequency and duration of muscle-strengthening activities^[44]^. Owing to the nature of the comprehensive items in the MSEQ Indonesian version, our pilot validity study found the higher validity of the MSEQ Indonesian version for assessing the weekly frequency and duration of MSE compared with the BRFSS in assessing whether participants met the recommended muscle-strengthening activities engagement at least twice per week (Cohen’s kappa range 0.40 to 0.52)^[42]^ and the CPS-3 in assessing the weekly duration of MSE (Spearman correlation of 0.71)^[43]^. The MSAS uses seven items to assess the frequency and duration of MSPAs, but no study has assessed its validity to date^[45]^.

The MSEQ Indonesian version showed better concurrent validity (r range from 0.30 to 0.36) against HGS than the available questionnaires, including the muscle-strengthening activity question of the European Health Interview Survey-Physical Activity Questionnaire and the total activity of the International Physical Activity Questionnaire-Long Form^[46]^. Therefore, the MSEQ Indonesian version could be utilized in examining the dose–response relationship of frequency, intensity, duration, and volume with muscle strength, which requires further epidemiological studies^[47],[48]^.

By considering the feedback and suggestions from end-users and experts from multidisciplinary teams, we translated and culturally adapted the MSEQ into its Indonesian version. The Indonesian version showed good validity and reliability for assessing the frequency, duration, intensity, volume, target muscle group, and different types of MSE. Although no gold standard is available for assessing all constructs of the MSE, we conducted construct validation studies using subjective and objective measures. In addition, we designed and reported our study under the COSMIN guidelines and provided detailed explanations of the terminologies used in our study to resolve several limitations of previous investigations, including poor methodologies and ambiguity of terminologies, as identified by a recent systematic review^[49]^.

### Limitations

The MSEQ Indonesian version could not cover all the dimensions of MSE, particularly repetition, number of sets, rest between periods, and load-based intensity^[50]^. However, we added duration into the MSEQ Indonesian version to allow for the analysis of the weekly volume of MSE. An additional limitation of our study was the representativeness of our respondents in the concurrent validity and reliability study. Although we recruited respondents from different types of universities and different geographical locations in Indonesia, they were not representative of Indonesian university students. Nevertheless, our study provided several important feasibility outcomes, including recruitment rates and dropout rates, which can be used in designing further large studies with representative respondents, either Indonesian university students or general Indonesian adults^[51]^.

## Conclusion

The MSEQ Indonesian version showed acceptable validity and reliability for assessing the weekly frequency, weekly duration, average intensity, weekly volume, and targeted muscle groups of MSE among university students. Further studies should be conducted by considering some of the feasibility outcomes reported in our study to examine the validity and reliability of the MSEQ Indonesian version among representative Indonesian university students and adults.

## Data Availability

All relevant data are within the manuscript and its Supporting Information files.

## Acknowledgements

We sincerely thank Universitas Gadjah Mada Publishing and Publication Agency for proofreading the manuscript. We also extend our gratitude to all experts and students who provided their input on the questionnaire.

## Contributors

RSA and RAW conceptualized and designed the study. RSA, DAP, and MNT collected the data. RAW and RMS monitored the data collection. RSA and MNT analyzed the qualitative data. RSA, STBM, and RAW analyzed the quantitative data. RSA drafted and revised the manuscript; RMS, STBM, DAP, MNT, and RAW wrote and revised the manuscript.

## Funding

This research was supported by the Junior Researcher Grant, Faculty of Medicine, Public Health, and Nursing, Universitas Gadjah Mada (Ref No:324 /UN1/FKKMK/PPKE/PT/2022).

## Competing interests

The authors declare that this research was conducted in the absence of any commercial or financial relationships that could be construed as a potential conflict of interest.

## Data availability statement

The anonymized collected data are available upon reasonable request from the corresponding author.

## Notes

### Competing Interest Statement

The authors have declared no competing interest.

### Funding Statement

Yes

### Author Declarations

This study obtained ethical approval from the Medical and Health Research Ethics Committee Faculty of Medicine, Public Health, and Nursing Universitas Gadjah Mada (Ref No: KE/FK/0973/EC). Informed consent from participants was obtained and documented before data collection.

